# Safety and efficacy of the Plasma Directed Electron Beam (PDEB™) - implications for enhanced wound healing treatment in military operational medicine and beyond

**DOI:** 10.1101/2024.02.21.24302399

**Authors:** Joseph A. Bauer, Adrianne R. Blocklin, Annette M. Sysel, Thomas J. Sheperak

## Abstract

**Introduction:** Wound healing presents a critical challenge in military operational medicine and combat casualty care, especially for soldiers in high-risk environments such as combat zones and training exercises. In these scenarios, wounds often result from bullets, shrapnel, burns, and blasts, affecting soft tissue, bone, and internal organs, and are frequently contaminated with hazardous substances like debris and bacteria. Limited resources in these environments make rapid and effective treatment difficult, often leading to delayed medical care and poorer healing outcomes. Emerging technologies like nonthermal plasma (NTP), also known as cold plasma, may provide superior wound healing treatment efficacy in these environments, owing to the ability to effectively kill pathogens, stimulate tissue regeneration, and minimize collateral damage compared to traditional methods. The Plasma Directed Electron Beam™ (PDEB™), an innovative advancement in nonthermal plasma research, shows promise in addressing these challenges.

**Materials and Methods:** The antibiofilm efficacy of the PDEB™ was investigated on Acinetobacter baumannii and Streptococcus mutans. Cytotoxicity was assessed using primary human epithelial cells and TR146 cells, immortalized epithelial cells. Cell proliferation assays, immunoblotting, and lactate dehydrogenase (LDH) release were evaluated.

**Results:** Our study demonstrates the effectiveness of the PDEB™ handheld in inhibiting the growth of bacterial pathogens implicated in biofilms. Acinetobacter baumannii and Streptococcus mutans showed zones of inhibition starting at lower power levels, achieving complete inhibition at 14 watts (W) and 7W respectively for 90-120 seconds. The safety of the PDEB™ was assessed through cell proliferation assays using human epithelial cells and semi-confluent TR146 cells, which were exposed to similar conditions as the bacterial assays. TR146 cells showed negligible differences in cleaved caspase 3 levels compared to controls. Cytotoxicity and apoptosis assays further confirmed the safety of PDEB™, as lactate dehydrogenase (LDH) release in epithelial cells and activated caspase 3 levels in cell extracts were comparable to untreated and helium-treated cells, indicating minimal cellular damage.

**Conclusion:** The PDEB™ handheld, a first-generation device, has demonstrated significant efficacy in inhibiting the growth of bacteria. Concurrently, its application on human epithelial cells has shown encouraging safety profiles. These findings align with the effectiveness of traditional nonthermal plasma devices, positioning the PDEB™ as a viable and promising option for wound healing applications in Combat Casualty Care and Military Operational Medicine.

## INTRODUCTION

Effective wound management is crucial in military medicine, disaster response, as well as remote and austere medicine, helping to save lives and mitigate the impact of medical emergencies in challenging environments. The U.S. military has made significant advancements in this area, notably through the Combat Wound Initiative Program and EMS training and manuals [1–3]. However, optimizing treatment for acute and chronic wounds remains a complex and multifaceted challenge that demands innovative solutions. The treatment of wounds involves various methods, including physical barriers like dressings and skin substitutes, which protect the wound, absorb excess fluid, and create a conducive healing environment [4]. Additionally, device-assisted healing, such as Negative Pressure Wound Therapy (NPWT), Modular Adaptive Electrotherapy Delivery System (MAEDS), hyperbaric oxygen therapy (HBOT), non-invasive wound closures and pharmaceutical interventions like antibiotics, antimicrobials, and blood products such as platelet-rich plasma (PRP) play vital roles in enhancing blood flow, stimulating tissue formation, and preventing infections [5–11]. Despite these advancements, a comprehensive solution for enhanced wound repair and chronic wound management remains elusive, moreover treatment modalities must be battlefield ready.

Nonthermal plasma (NTP) have been sought after to augment or replace conventional wound healing therapies [12]. The value-added proposition of the PDEB™ is its ability to disrupt bacterial biofilms using a site-directed beam of electrons and activation of the apoptotic cascade of cell death by nitric oxide and reactive oxygen and nitrogen species (RONS) and within surrounding tissues [13]. RONS initiate a cascade of apoptotic events that contribute to the disruption of biofilms and eradication of bacteria far below the tissue surface [14, 15].

Nitric Oxide (NO) is a well-studied intercellular signaling molecule, is involved in key biological processes that help wound healing, namely angiogenesis, inflammation, and collagen deposition [16–21]. NO is vital to the activity of proangiogenic cytokines, such as VEGF, that can then stimulate the formation of new blood vessels to help in wound healing [18]. It can also modulate chemo-attractants that promote the infiltration of neutrophils and monocytes to the wound site. Once these cells arrive, they release TNFα and IL-1, which in turn ushers in the keratinocytes to reestablish the epithelial barrier that was disrupted during the injury [20, 22]. NO promotes the synthesis and deposition of collagen by fibroblasts, which then helps to strengthen the extracellular matrix and accelerate the wound healing process [16, 17].

NO also plays an essential role as an antibacterial agent by reacting with oxygen and superoxide to form dinitrogen trioxide (kills by deaminating DNA) and peroxynitrite (kills by lipid peroxidation and membrane damage) [21, 23, 24].

However, abnormally low production of NO has been linked to impaired wound healing and the development of chronic wounds [25]. Serum levels of NO may also be used as prognostic markers, with abnormal NO concentrations serving as reliable predictors of poor healing outcomes in septic burn patients and chronic diabetic foot ulcers (DFUs) [26, 27]

Current evidence suggests that diabetic wound fluid has significantly lower levels of NO than healthy wound fluid, due to downregulated endothelial nitric oxide synthase (eNOS) [28, 29]. Macrophages, keratinocytes, and fibroblasts all express elevated levels of inducible nitric oxide synthase (iNOS) at a healthy wound site, but that expression is suppressed in diabetes. In the absence of the recruiting effect of NO, macrophages are found in lower densities in chronic wounds, diminishing pathogen clearance [30, 31]. Inhibited NO production strongly reduces the number of keratinocytes during the process of re-epithelization and further antagonizes keratinocyte proliferation and differentiation [32–34]. Fibroblasts in diabetic chronic wounds express lower levels of both iNOS and eNOS, become senescent, and do not produce collagen or form the extracellular matrix (ECM). Thus, decreased NO in extracellular wound fluid corresponds with decreased collagen content and weaker wound breaking strength [35].

Nonthermal plasma technology offers significant promise in wound healing in large part due to its ability to produce NO. However, its use is limited due to the production of potentially harmful byproducts [36]. This limitation underscores the need for refined methods to effectively harness the therapeutic potential of nitric oxide. Addressing this, a novel approach is proposed using nonthermal plasma technology to deliver therapeutic levels of nitric oxide without undesirable byproducts, thus overcoming the challenges associated with nitric oxide’s unstable nature and delivery. Proof of concept experiments were conducted with the PDEB™ handheld prototype to evaluate safety and efficacy as an early step leading to the development of a device designed for tactical combat casualty care and enhanced wound healing outcomes.

## MATERIALS AND METHODS

### Cell Culture

Cell lines TR146 cells, immortalized epithelial cells were obtained from the American Type Culture Collection (ATCC, Manassas, VA). Primary human epithelial cells were obtained from biopsy according to protocols approved by the Institutional Review Board of Case Western Reserve University as previously described [37]. Cells were maintained according to ATCC instructions. Cells were maintained in 5% CO_2_ at 37°C in a humidified tissue culture incubator. Cell lines were routinely checked for mycoplasma contamination and were found to be mycoplasma free.

### Bacterial Assays

Bacteria strains Acinetobacter baumannii (AB0057) and Streptococcus mutans (25175) were purchased from American Type Culture Collection and inoculated on agar plates according to manufacturer’s instructions using Mueller– Hinton (MH) medium (MilliporeSigma; Burlington, MA). An individual colony was selected and grown to midlog phase, in accordance with standard microbiological techniques, by growing the culture in MH medium overnight at 37°C and 275 rpm in a shaking incubator.

### Cell Proliferation Assay

Cells were exposed to the PDEB™ for various intensities and duration. Cell counting was performed using the Crystal Violet staining method. Cells were seeded in 96-well plates and allowed to adhere overnight at 37°C in a 5% CO_2_ atmosphere. Following the treatment period, cells were fixed with 4% paraformaldehyde for 15 minutes at room temperature. After fixation, cells were stained with 0.1% Crystal Violet solution for 20 minutes. Excess stain was washed off with distilled water, and the plates were air-dried. The dye was solubilized in 10% acetic acid, and the absorbance was measured at 590 nm using a microplate reader.

### Gel Electrophoresis and Immunoblot analysis

Cells were challenged at various time points and intensities with the PDEB™ in 96 well plates. Whole cell lysates were prepared in a high salt lysis buffer for subsequent immunoblotting studies. Total cell protein was resolved by electrophoresis and transferred to a membrane according to standard molecular biology methods. The primary antibodies were incubated overnight: mouse anti-cleaved caspase 3 (1:500, Pierce, Rochford IL); or mouse anti-caspase 3 (1:1000, Pierce, Rochford IL). Membranes were washed and incubated with horseradish peroxidase-conjugated goat secondary (1:2000, BioRad, Hercules, CA). The protein bands were visualized by chemiluminescence. Antibodies against glyceraldehyde-3-phosphate dehydrogenase (mouse anti-GAPDH, 1:3000, Proteintech, Rosemont, IL) were used to ensure equal loading.

### Lactate Dehydrogenase Assay

Cytotoxicity assays were performed on PDEB™ challenged samples using the Cytotoxicity detection kit (Millipore Sigma, Burlington, Massachusetts) according to manufacturer’s instructions.

### Gas Chromatography Mass Spectroscopy Analysis

Samples from the PDEB™ were obtained directly from the generator’s output into a gas tight syringe (Hamilton Company, Reno NV) and injected into a mass spectrometer. Shimadzu GC-MS Spectrometer QP2010 Plus was equipped with molecular sieve column Agilent CP-Molsieve 5Å (25 m length, 0.32 mm diameter, 30 μm film thickness). Shimadzu GC-MS QP5050A instrument with a polysiloxane column (Phenomenex ZB1-MS, 30 m length, 0.25 mm diameter, 0.25 μm film thickness). The instrument operated in the SCAN mode recording signal from all the possible ions in the range 20-100.

## RESULTS

### PDEB™ produces pure on-demand NO

The PDEB™ gas stream was drawn directly from the generator’s output into a gastight syringe and injected into the mass spectrometer. The Shimadzu QP2010 GCMS was equipped with a molecular sieve column which separated nitric oxide (NO) from the air components: O_2_, N_2_ and argon. Using Selected Ion Monitoring (SIM) mode, the presence of ions characteristic for nitric oxide (30 molecular weight; MW), nitrogen dioxide (46 MW) and ozone (48 MW) were assessed (Figure 1). The nitric oxide peak was visible and there were no other peaks shown. Additionally, we evaluated a polysiloxane column to mitigate concerns of possible NO_2_ retention by the Molsieve column[38].

**Figure 1.**
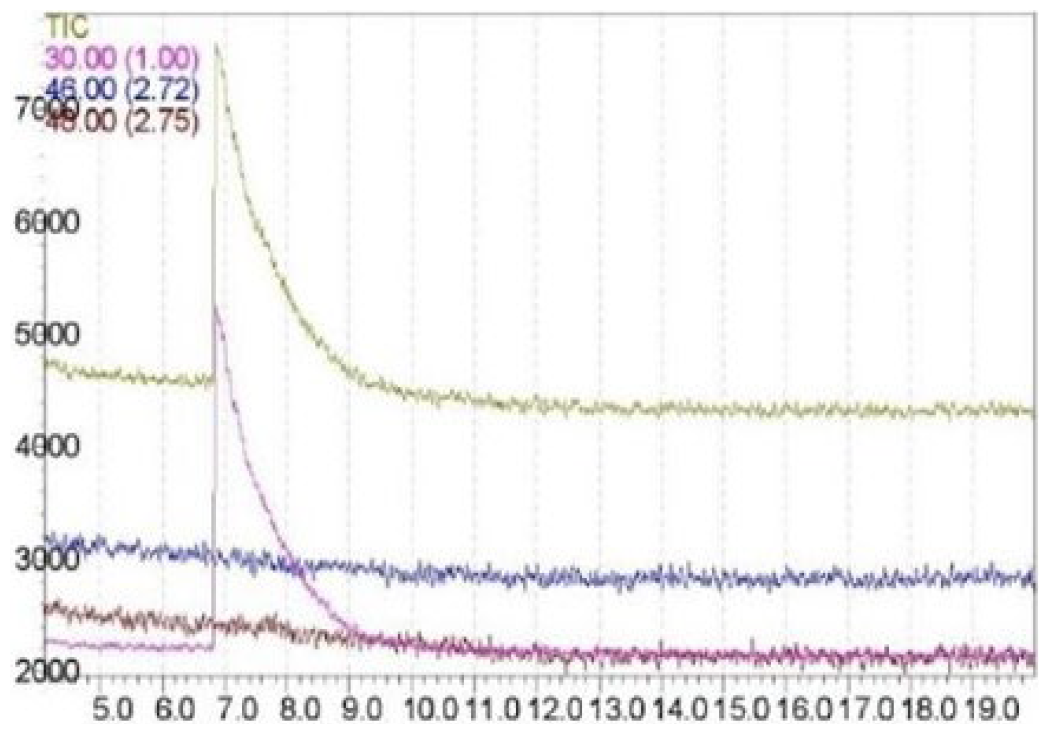
Select Ions Monitoring (SIM) Chromatograms for NO (30), NO_2_ (46), O_3_ (48) and the Total Ion Chromatogram (TIC) from a PDEB™ Output Stream Gas Aliquot.

The analysis was repeated using a Shimadzu QP5050A GCMS instrument with a polysiloxane column. The instrument operated in the SCAN mode recording signal from all the possible ions in the range of 20-100 MW. The signal from the ion of the mass 30 MW (nitric oxide) was strong while 46 MW (nitrogen dioxide) and 48 MW (ozone) were absent (data not shown). This confirms that nitrogen dioxide and ozone are not present as ozone and nitric oxide would lead to formation of nitrogen dioxide. Thus, the PDEB™ has been shown to produce pure NO under these testing conditions.

### PDEB™ anti-bacterial scouting experiments

PDEB™ was used to conduct proof of principle experiments showing that the unit could inhibit the growth of bacteria while maintaining viability of human epithelial cells; the treatment is shown in Figure 2A. Acinetobacter baumannii was challenged with the plasma beam at a distance of 3 mm from the agar surface, for various times (in seconds) and intensities (5W-8W) (Figure 2B). Notice zones of inhibition that can be seen as early at 5W, 120,” and culminating with inhibition through the entire width of the agar at 7W, 120”. Streptococcus mutans was challenged with the plasma beam for various times (in seconds) and intensities (8W-14W) (Figure 2C), at 3 mm from the agar surface. Notice zones of inhibition that can be seen as early at 8W, 90”, and inhibition is observed through the entire width of the agar at 14W, 90”.

**Figure 2.**
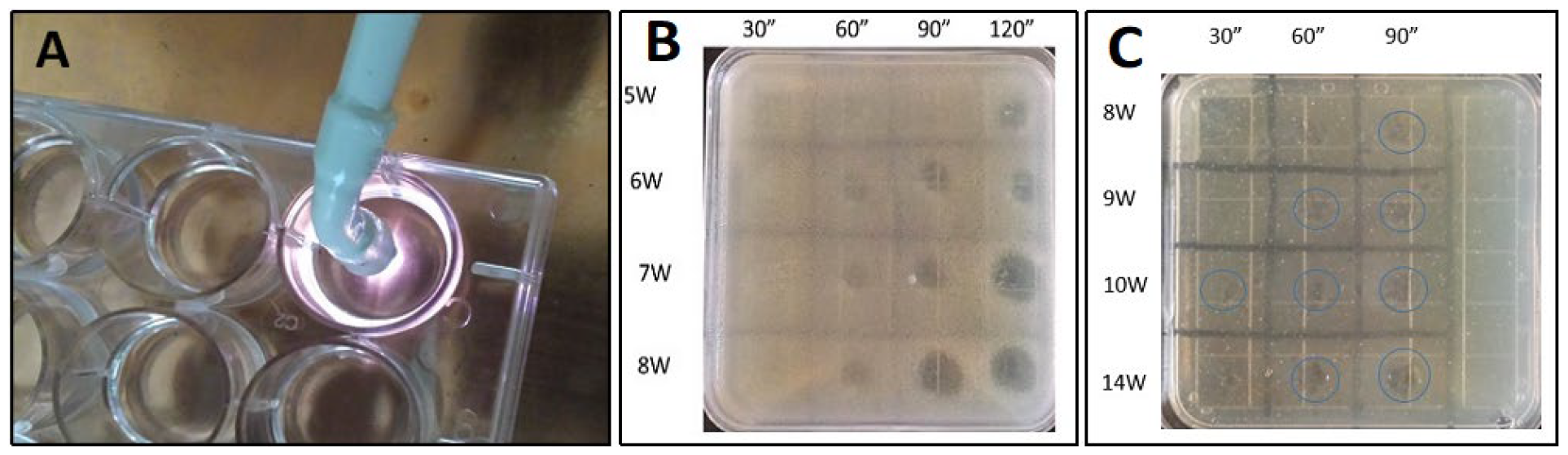
[A] NTP treatment of a multi-well plate. NTP challenge of A. baumannii [B] and S. mutans [C] in pour plate agar.

### PDEB™ safety studies against normal human cells

Preliminary safety experiments were conducted using human epithelial cells that were challenged with the PDEB™ under similar conditions described for the bacterial inhibition assays (Figure 3).

**Figure 3.**
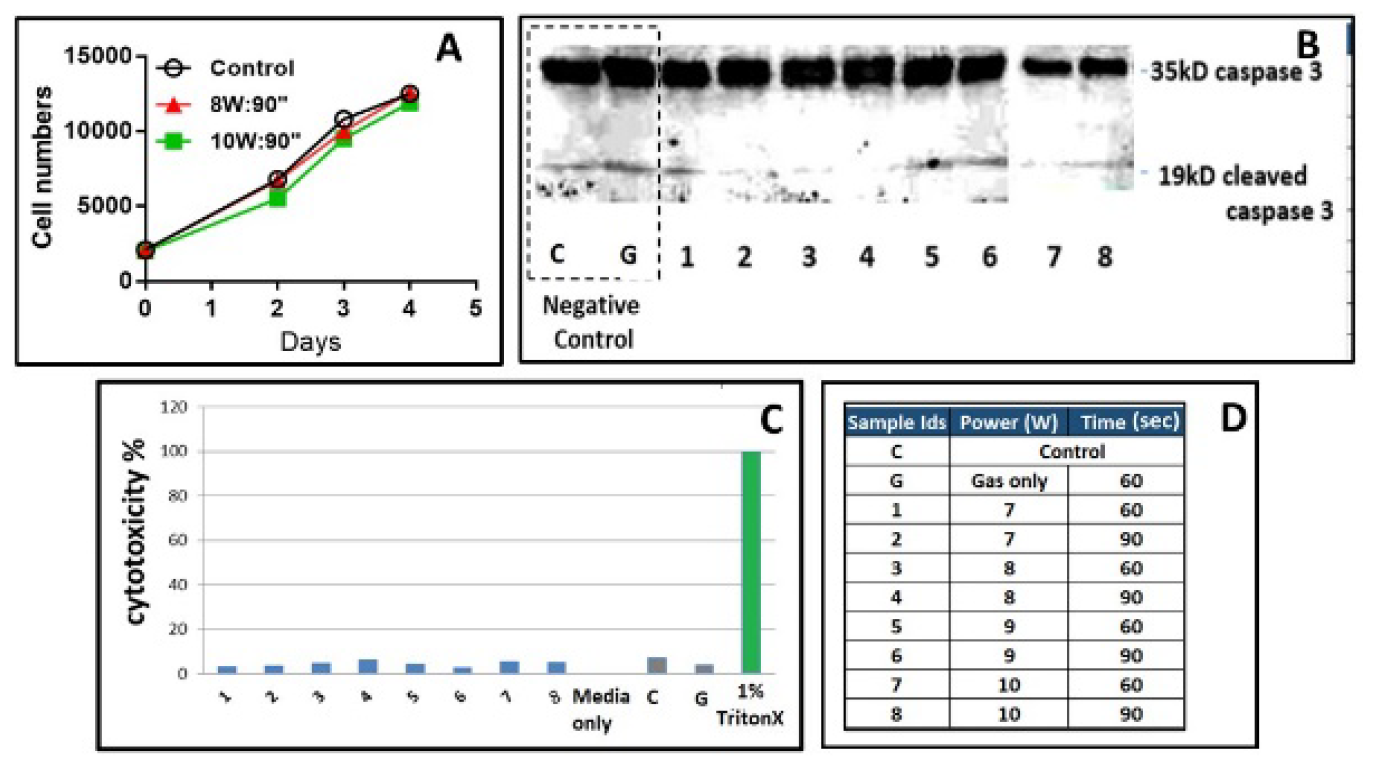
[A] Cell proliferation curves of plasma challenged TR146 epithelial cells [B] Western blot analysis assessing apoptosis in epithelial cells challenged with plasma. [C] Plasma challenge of epithelial cells followed by LDH assay to determine cytotoxicity. [D] conditions (intensity and time) used in the LDH assay.

TR146 cells (immortalized epithelial cells) were grown in complete medium until semi-confluent (80%). They were then challenged by the plasma beam (1 mm distance, at respective intensities and times). Following trypsinization, the cells were loaded into 96 well plates (40,000 cells per well). Following respective days of growth, post-plasma challenge, cells were detached via trypsin and counted (Figure 3A).

Cellular extracts from the proliferation assay were run on SDS-PAGE, followed by Western blotting, to determine cleaved caspase 3 (activated caspase; “the executioner caspase” that promotes apoptosis) (Figure 3B). Sample numbers and plasma conditions are the same as those listed in Figure 3. Notice that when compared to the two negative controls (C, cells alone; G, cells exposed to gas), the level of cleaved caspase 3 was negligible in all the conditions. Primary human epithelial cells were challenged with NTP and the percentage of lactate dehydrogenase release in respective media was determined (Figure 3C and D). Notice that the levels of LDH in respective media were negligible, and comparable to that of C (cells alone) and G (cells challenged with gas [industrial grade helium]).

## DISCUSSION

Our PDEB™ handheld prototype was shown to inhibit the growth of various bacteria inherent in biofilms including Acinetobacter baumannii (multi-resistant) and Streptococcus mutans. Furthermore, studies of this technology with human oral epithelial cells provide promising indications for safety. These results are consistent with the reported efficacy of traditional nonthermal plasma devices and establish PDEB™ technology as a promising treatment modality for wound healing in austere and threatening environments [39].

Nitric oxide (NO) represents a potential wound therapeutic agent due to its ability to regulate inflammation and eradicate bacterial infections. Many studies support the utility of NO as a therapeutic agent for wound healing [40]. The use of NO as a wound healing therapeutic may be categorized by two broad strategies: generating NO from endogenous reservoirs, and dosing with NO from exogenous sources. Generation of NO from endogenous sources may be accomplished by L-arginine supplementation as a substrate for iNOS [41]. Dosing exogenous NO can be accomplished by treating wounds using gaseous NO or formulations of nitrite under acidified conditions. Nitrite and nitrate are known precursors of NO [41, 42]. Further, small molecule NO-releasing donors can be applied directly to wounds encapsulated via polymers, gels, and/or dressings [43–45]. Nitric oxide donors can also be chemically attached to macromolecular scaffolds, providing potentially greater stability to labile NO donors and enabling targeted, tunable delivery of NO [46]. The goal of both endogenous and exogenous therapeutic NO dosage is to regulate inflammation and tissue remodeling to promote wound healing.

Gaseous NO as a therapeutic agent for non-healing wounds has also been explored in terms of antimicrobial action. Studies have reported that gaseous NO could be safely dosed for 8 h at levels of 5, 25, 75, and 200 ppm [47]. None of these studied gaseous NO concentrations damaged the extracellular matrix, some even increased lymphocyte proliferation. These results suggest that wound infections can be treated with NO at concentrations of 5–200 ppm. Higher concentrations of NO (up to 500 ppm) have also been tested but for only 60 seconds once-daily for 6 days to mitigate toxicity concerns [48].

The therapeutic potential of gaseous NO is often limited to a hospital setting due to the hazards associated with pressurized NO cylinders and the need for continuous oversight [49]. Additional concerns regarding gaseous NO delivery include NO’s high reactivity, particularly with oxygen in the air to form harmful byproducts, such as nitrogen dioxide (NO_2_) [50]. This reactivity often necessitates that gaseous NO therapy occur in anaerobic environments, limiting the potential for therapeutic use outside of a hospital setting [51]. Both NO and NO_2_ pose serious systemic toxicity concerns when inhaled, starting at concentrations as low as 40 and 1.5 ppm, respectively [52]. Thus, significant safety protocols and monitoring are required for gaseous NO treatments of wounds, which limits its usefulness.

The key difference of the PDEB™ compared to conventional NTP devices is its ability to produce a focused beam of electrons and pure nitric oxide, an innovative benchmark for the field. This is in contrast to a major drawback in traditional NTP devices; the indiscriminate production of gases including NO, NO_2_, H_2_O_2_ and O_3_ which are toxic at the levels generated [53]. The ability of the PDEB™ to generate NO gas on-demand (without the use of high-pressure nitric oxide gas cylinders) adds to the safety of the device. Compressed NO gas is extremely dangerous and an immediate health hazard. The rate of conversion from NO to NO_2_ is directly proportional to the pressure of the cylinder. Compressed NO gas (500 PSIG) exposed to air reacts immediately to produce NO_2_ for which the OSHA Permissible Exposure Limit-Ceiling (PEL-C) is 5 ppm). The PDEB™ produces NO on-demand, under ambient conditions, which does not result in NO_2_ levels that exceed OSHA safety standards. [50, 52].

Nitric oxide, recognized for its critical role in wound healing, offers significant promise in the context of combat casualty care. Among the technologies harnessing the power of NO, the PDEB™ is novel for its on-demand NO production from a handheld sized device. The device is distinct in its ability to produce a focused beam of electrons and pure nitric oxide, a marked improvement over traditional nonthermal plasma devices which often generate a mix of gases. The PDEB™’s capability to generate NO gas under ambient conditions is particularly advantageous in the challenging environments of combat casualty care. It circumvents the risks associated with compressed NO gas, which is not only hazardous but also impractical in field conditions. The on-demand generation of NO at the site of injury can offer immediate therapeutic benefits, enhancing wound healing processes while minimizing the risk of infection, a crucial aspect in combat injuries whereby delayed or inadequate treatment can lead to severe complications. Moreover, the device’s design aligns with the logistical and safety requirements of battlefield scenarios, offering a portable, efficient, and safer alternative to traditional wound healing methods. This alignment with the needs of combat casualty care positions the PDEB™ as a potentially transformative tool in the management and expedited healing of wounds in such critical and urgent environments. Future studies will focus on a lightweight, portable, battery-operated device for individual or EMS/medic use.

## Data Availability

All data produced in the present study are available upon reasonable request to the authors.

## ACKNOWLEDGEMENTS

A very special thanks to Case Western Reserve University, Department of Biological Sciences for performing the safety and efficacy experiments with the assistance of Thomas Sheperak. Special thanks to Jedrzej Romanowicz of Bowling Green State University, Department of Chemistry for conducting the GC-MS experiments.

